# VideoCap: Enabling REDCap as a Tool for Crowdsourced Multimedia-Labeling Research

**DOI:** 10.64898/2026.06.26.26355995

**Authors:** Basam Alasaly, Kuk Jin Jang, Andrew Zolensky, Sriharsha Mopidevi, Kevin B. Johnson

**Affiliations:** Division of Informatics, Department of Biostatistics, Epidemiology, and Informatics, Perelman School of Medicine, University of Pennsylvania, Philadelphia, PA, USA; Department of Computer Engineering, Hongik University, Seoul, South Korea; Graduate Group in Genomics and Computational Biology, Perelman School of Medicine, University of Pennsylvania, Philadelphia, PA, USA; Department of Computer and Information Science, School of Engineering and Applied Science, University of Pennsylvania, Philadelphia, PA, USA

## Abstract

**Objective:** Machine learning systems that use video data require large, diverse, human-labeled datasets, but generating reliable annotations remains labor-intensive, difficult to scale, and often dependent on proprietary tools or small expert annotator pools. We present VideoCap, a secure and context-aware workflow that integrates REDCap with a Content Delivery Network (CDN), backend web service, and crowdsourcing platform to dynamically rotate embedded video segments within a single survey structure.

**Methods:** VideoCap hosts segmented videos on a CDN with restricted downloads, stores segment metadata and session state for automated video selection, and generates REDCap survey URLs populated with embedded video parameters. We implemented the workflow through Amazon Mechanical Turk to annotate simulated patient-provider video segments using open-ended insights, structured scheme selections, and free-text responses.

**Results:** We tested the workflow using 481 simulated patient-provider video segments over a 131-day deployment period. In the valid-only analytic subset, 358 unique video segments received 814 annotations, with an average of 2.27 annotations per segment. The workflow achieved an average annotation-time-to-video-duration ratio of 4.47:1, lower than contextual annotation-time estimates reported in prior multimedia annotation workflows.

**Conclusion:** VideoCap provides a reproducible workflow for dynamic multimedia annotation using broadly accessible tools, demonstrating feasible survey delivery and efficient annotation.

## Introduction

Machine learning systems for visual data depend on large, diverse labeled datasets, and model performance has been shown to improve as training data volume increases.(1) For video-based research, producing those labels remains a central bottleneck because each usable annotation requires human time, judgment, and repeated exposure to multimedia content. The demand for labeled video continues to rise across surveillance, education, social media, and clinical domains, where roughly 720,000 hours of video reach a single platform daily.(2) The constraint sits in the annotation step, where human capacity governs how much of this video becomes usable training data.(3)

The standard solution requires domain experts to perform that annotation. Experts deliver precision, but their scarcity limits throughput and diversity. A small pool of specialists carries knowledge and perspectives that diverge from the general population, which can yield labels shaped by that narrow vantage.(4) Models trained on such data may perform well in controlled conditions and degrade in real-world settings.(5) Broadening the respondent pool may help restore the diversity these models need.(6)

Existing video annotation and summarization methods inherit these limits. Many segment video too finely or too coarsely and lose the context of events, and most run on proprietary, in-house tools that resist reuse.(7–12) These approaches also overlook the spatiotemporal relationships that connect events, which can produce annotations that lack coherence and narrow their usability.(3,13)

An annotation workflow built on established, broadly accessible survey software would remove these reuse barriers and support deployment across a wider respondent pool. REDCap (Research Electronic Data Capture) is an established software application developed at Vanderbilt University to support data collection and management for clinical and translational research. Since its inception, REDCap has grown significantly, with 7,000 partner institutions across 163 countries, making it a widely adopted tool in global research that supports thousands of studies across various domains.(14) However, REDCap is limited in supporting large multimedia library annotation because its primary function consists of the creation of static survey structures, and it does not allow for dynamic integration of multimedia content into that structure. It also does not have built-in Content Delivery Network (CDN) capabilities, making it difficult to manage and distribute large files efficiently.

To close these gaps, we developed VideoCap, a reproducible workflow that extends REDCap to support mass human annotation of large-scale multimedia datasets using existing survey software. We aim to provide crowdsourced respondents with a single survey URL in which a multimedia placeholder is populated with content from a CDN-hosted library that advances sequentially with each survey access, independent of respondent identity. This solution is cost-effective, as the major burden of hosting the survey software is handled by institutions. By using a single access point that does not rely on respondent identity, this solution can be released to multiple crowdsourcing platforms simultaneously. Respondent and platform identity are maintained via a survey-generated completion code submitted on the crowdsourcing platform.

## Related Works

Prior large-scale video datasets show a dominant annotation pattern in this space: project-specific pipelines built around expert or professional annotators, custom-built tools, and dataset-specific quality-control procedures. These workflows are often difficult to reproduce, adapt, or redeploy across institutions and domains. The Kinetics dataset demonstrates this pattern: although it scaled to approximately 650,000 video clips across 700 action classes, its annotation process relied on expert annotators using custom-built tools to ensure high-quality labels.(7,9) The AVA dataset, introduced by Google Research, which focused on spatiotemporal localization with detailed frame-level annotations of 80 atomic actions, also used professional annotators and specialized tools.(8) The AViD dataset, by Piergiovanni et al., which aimed to reduce cultural biases by including videos from diverse regions, combined automated labeling with manual verification; however, it still required custom-developed tools.(11) The BDD100K dataset, by Yu et al., which was designed for autonomous driving research with 100,000 videos and comprehensive annotations, utilized skilled annotators and custom tools specific to driving scenarios; again, limiting the transferability and accessibility of its methods to other domains and use cases.(12)

Scalability remains a major challenge due to the vast volume of data, the need for proprietary tools, and the reliance on expert annotators, thus limiting widespread applicability.(7,8) The lack of accessible, adaptable multimedia annotation processes hinders progress, as custom tools developed for specific datasets are often not publicly available, nor are they intuitive to a new audience of annotators.(8,12) Our work addresses these challenges by introducing a modular crowdsourced multimedia annotation framework that leverages reputable and accessible industry-standard tools.

Our modular workflow is reproducible and adaptable across domains, addressing limitations of existing methods that are built custom for specific applications. By supporting crowdsourced annotation through broadly accessible tools, VideoCap may help researchers collect larger and more diverse multimedia annotation sets while using quality-control mechanisms, including worker qualification requirements and periodic response monitoring, to help maintain annotation standards despite the inherent variability of crowdsourced labor.(10)

## Methods

We developed a scalable workflow for video annotation that leveraged REDCap, video segmentation and hosting, and backend functions to provide efficient, respondent-specific video delivery while maintaining content security, as shown in Figure 1.

**Figure 1.**
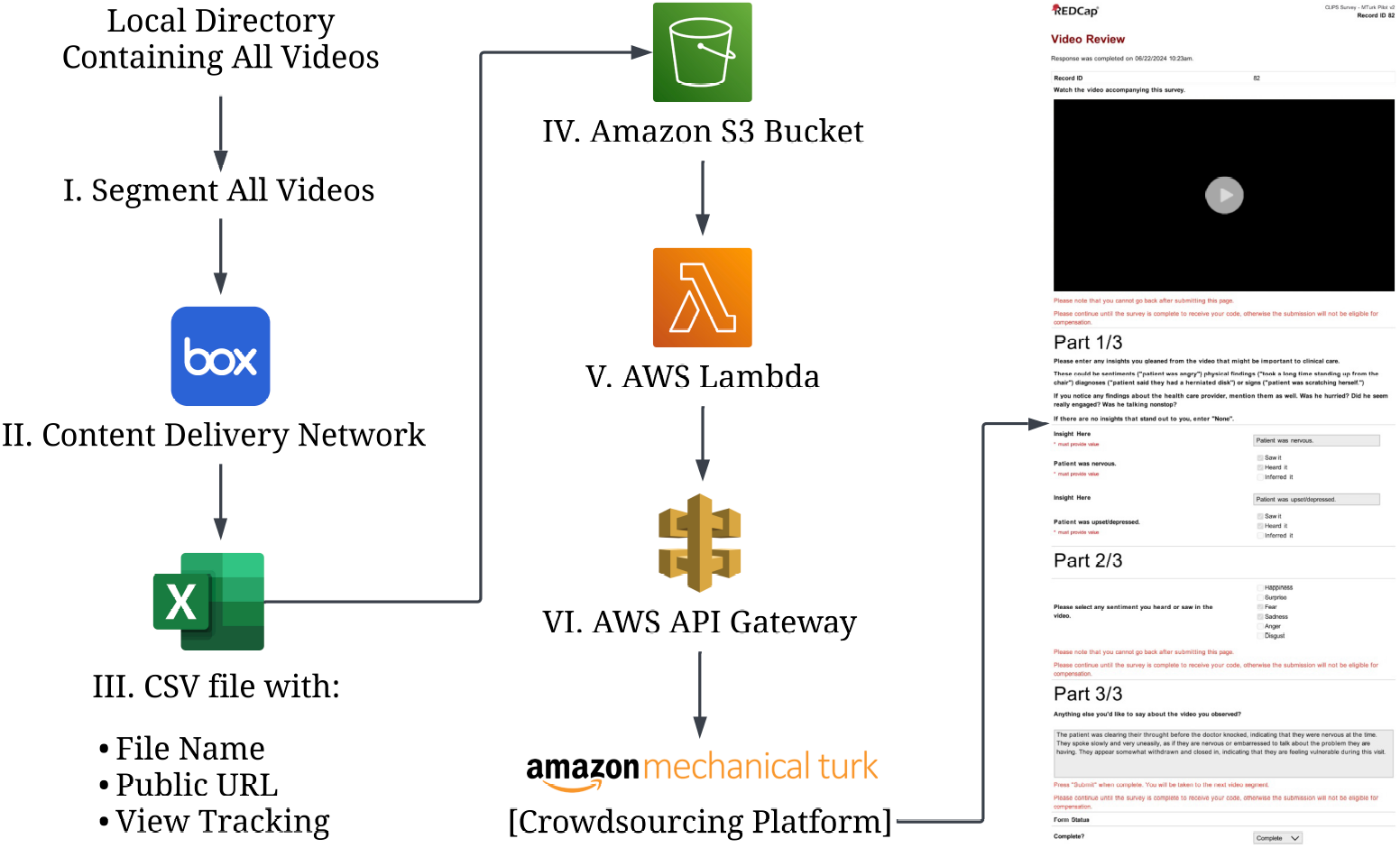
Workflow diagram illustrating the main steps in the video annotation process. Icons sourced from Seeklogo and Creazilla, with some modifications.

Our workflow incorporated several critical design considerations:

- A single, universal access point for respondents that generated unique survey URLs with automated video selection
- Dynamic video embedding within the survey that allowed for flexible content delivery without creating multiple survey versions
- Restricted content download for sensitive data

### Workflow Description

The workflow comprises six core stages that carry a set of videos from media processing through dynamic REDCap survey-link generation. We then describe how the workflow was deployed on a crowdsourcing platform for annotation.

#### Core Workflow Components

Stages I-VI describe the reusable VideoCap infrastructure used to segment media, host content, catalog embedded URLs, maintain session state, and generate dynamic REDCap survey links.

##### I. Video Segmentation

Given a set of videos, to make the annotation tasks manageable, we divided each video into 1-minute segments. If the final segment was shorter than 30 seconds, it was appended to the previous, ensuring that the last segment did not exceed 1 minute and 29 seconds.

##### II. Content Delivery Network

The segmented videos were hosted on Box, Inc., a file sharing tool and Content Delivery Network (CDN) that supports embedding within REDCap and restricts video access to view-only to safeguard against unauthorized content download. This choice allowed for efficient and secure distribution of the media content to respondents.

##### III. Video Metadata Management

After segmentation and content upload to Box, we generated a comma-separated values (CSV) file that organized each video segment’s name and corresponding public URL. A Python script was used with Box’s application programming interface (API) to batch-generate all embeddable segment URLs and create the CSV file in a single operation. The CSV file included:

- File Name
- Public URL
- View Tracking

##### IV. Object Storage: Media Catalog and Session State

Object storage holds the media URL list and session state required for dynamic survey delivery. In our implementation, we used Amazon Simple Storage Service (S3) to store a CSV file containing each video segment’s name, embeddable URL, and view-tracking fields. A separate text file tracked the current position in the media sequence, including the video index and round index. The backend service used these files to select the next segment or batch of segments, update viewing status, and continue the rotation when the end of the media library was reached.

##### V. Serverless Function

A serverless function drives the dynamic video delivery system, selecting and batching video segments for each session according to a predefined sequence. We implemented it in AWS Lambda, which executes code in response to defined triggers without provisioning or managing servers. The function performed the following tasks:

- **Data Retrieval and Parsing**: Retrieved video metadata from the CSV file stored in Amazon S3 and organized video segments by sequence.
- **State Management**: Maintained a state file in Amazon S3 to track the current position in the video sequence, including the video index and round index, and ensured that each successive respondent received unique videos. The state file enabled efficient batch handling by grouping four video segments per session.
- **Batching and Round-Based Viewing**: Selected a batch of four videos for each session, marked them as viewed in the CSV file, and advanced the state to the next batch upon completion. If the video index reached the end of the list, the round index was incremented to support multiple review rounds.
- **Dynamic URL Generation**: Generated a REDCap survey URL with video-specific parameters and a unique, randomized session ID. This allowed for response tracking between the survey results, the crowdsourcing platform, and the respondent, facilitating verification of response completion, origin, and quality.
- **Automatic Survey Redirection**: Created a redirect page that automatically directed users to the generated REDCap survey URL with the appropriate video parameters.

##### VI. API Gateway

An API gateway served as a single access point URL to the Lambda function. When a crowdsourcing worker accessed this URL, API Gateway called the Lambda function, which then redirected the user to the dynamically generated REDCap survey URL with respondent-specific video segments. This setup ensured that each respondent was seamlessly directed to a unique video set within the same survey structure.

#### Survey Deployment and Evaluation

The remaining sections describe how the core workflow was deployed for this study using a crowdsourcing platform and a REDCap survey instrument.

#### Crowdsourcing Platform Integration

The URL generated via the gateway was distributed to Amazon Mechanical Turk workers. This single access point allowed us to host one task on the crowdsourcing platform that facilitated annotation across a vast library of content, where each worker received a unique set of videos for annotation.

#### Survey Structure and URL Parameterization in REDCap

To support efficient video annotation, we structured the REDCap survey to dynamically embed video segments using URL parameters generated by the AWS Lambda function. By utilizing the URL parameter feature in REDCap, we passed each video segment into the survey and embedded each segment as a unique URL variable. We configured the parameters on the survey’s first page, hiding these unnecessary input fields from users with the @HIDDEN action tag to reduce distractions and help participants focus on the survey instructions and video content.

#### Survey Implementation and Evaluation

We implemented this workflow to annotate a set of 481 simulated patient-provider video segments. The resulting survey was deployed to Amazon Mechanical Turk workers to annotate the segments with key insights regarding the interaction between the patient and their provider.

We present the survey implementation and evaluate workflow deployment, annotation coverage, and efficiency, including the Annotation Burden Factor (the ratio of annotation time to video duration).

## Results

### Workflow Deployment and Survey Delivery

Our REDCap survey consisted of up to eight video segment reviews, each comprising a single output-only viewing component followed by three subsequent input components, shown in Figure 2, per segment.

**Figure 2.**
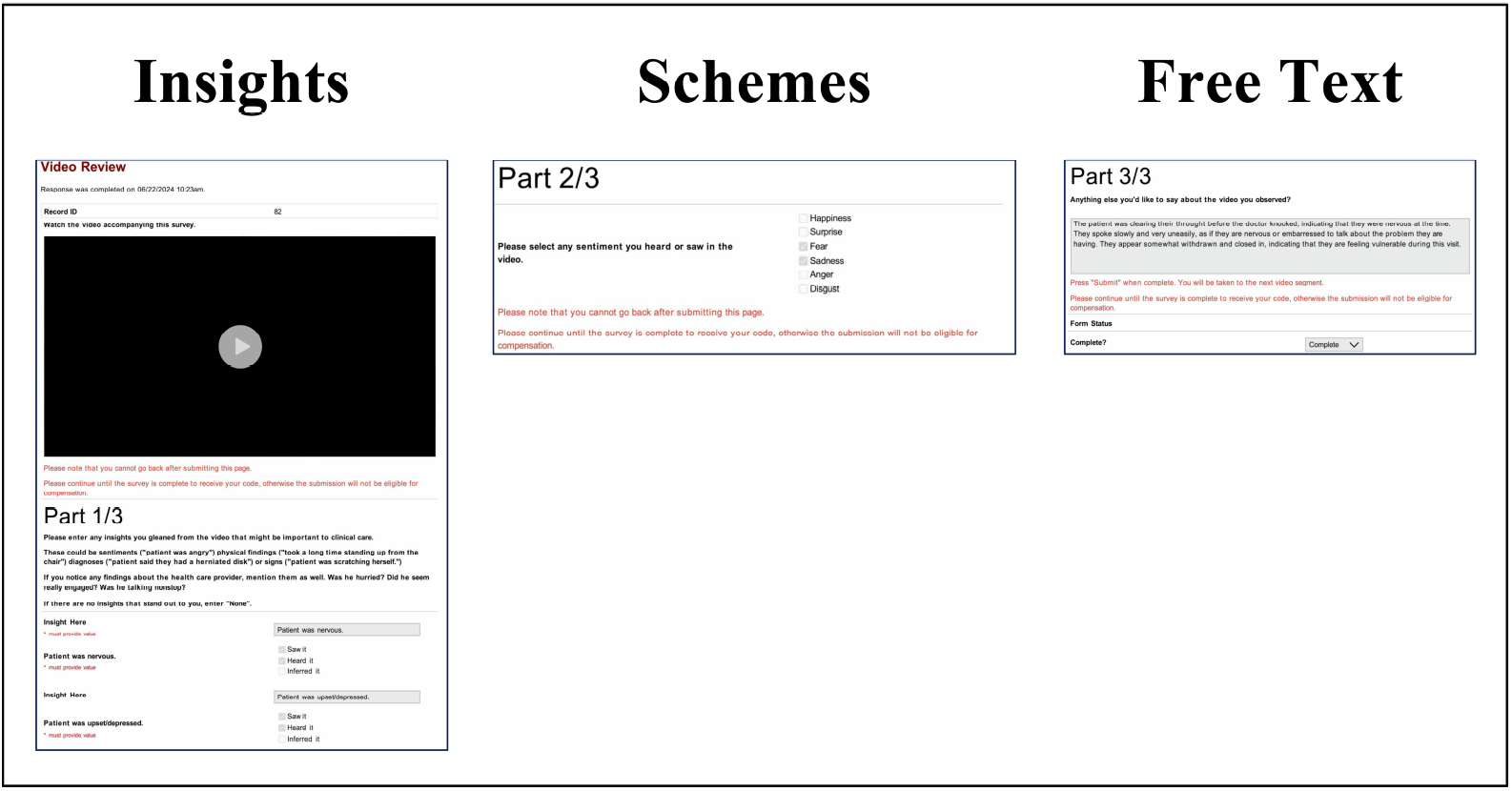
REDCap survey video review process. Each assigned video segment was presented as an output-only viewing component followed by open-ended insights, structured scheme selection, and free-text response fields.

The output component displayed the assigned video segment and asked participants to view it with no input allowed. The following insights view requested that the user enter up to five short open-ended insights, or “none,” if they believed that none existed. The schemes view then asked the user to select from a structured set of predefined schemes. Finally, the free text view was completely open-ended and invited the user to add any additional comments.

The workflow was deployed using a single access point distributed through Amazon Mechanical Turk. This access point directed respondents to dynamically generated REDCap survey URLs populated with assigned video parameters, allowing one REDCap survey structure to support annotation across the video library.

### Annotation Efficiency

Table 1 summarizes the workflow deployment and annotation efficiency. The workflow delivered 481 video segments for annotation. The valid analytic subset comprised 814 annotations across 358 video segments, corresponding to a mean of 2.27 valid annotations per segment.

**Table 1.** VideoCap Annotation Coverage and Efficiency Metrics.

| Metric | Value |
| --- | --- |
| Video segments available for annotation | 481 |
| Video segments with valid annotations, n (%) | 358 (74.4%) |
| Valid annotations | 814 |
| Mean valid annotations per annotated segment | 2.27 |
| Total annotation time | 60.7 h |
| Mean annotation time per response | 268.34 s (4.47 min) |
| Mean video duration | 60 s (1 min) |
| Annotation Burden Factor* | 4.47 |

\* Annotation Burden Factor was calculated as mean annotation time per response divided by mean video duration (268.34 s / 60 s).

Analysis of valid responses resulted in an average annotation time of 268.34 seconds for a mean video duration of 60 seconds. This yields an Annotation Burden Factor of 4.47, indicating that for every minute of video footage, crowd workers required approximately 4.5 minutes to view the content, interpret the context, and complete the multi-step classification tasks.

## Discussion

Our proposed modular workflow extends the capabilities of REDCap to support scalable and secure multimedia annotation processes with a flexible, adaptable design. Although we utilize AWS Lambda, S3, and API Gateway, the architecture is designed to be agnostic. Alternative backend services that offer similar functionality can be used instead. Likewise, any survey software that supports URL parameters can replace REDCap, as long as it works with a compatible Content Delivery Network (CDN) for secure media embedding and mass embeddable URL generation to avoid labor-intensive setup. This modular structure allows for adaptability across various systems while maintaining annotation quality, scalability, and security.

Individual components of VideoCap may also be reused outside the full crowdsourcing workflow. For example, the CDN-hosting and URL-generation steps can support smaller expert-review studies in which REDCap survey links are distributed manually, while the complete VideoCap workflow adds backend routing, sequential media rotation, and crowdsourcing-platform integration for larger-scale annotation.

Existing media annotation methods often struggle with scalability and consistency when dealing with extensive datasets.(7–9,11,12) By contrast, using VideoCap enabled us to distribute the workload across a broad pool of annotators. Integrated with REDCap and AWS, this solution is cost-effective and accessible, removing the need for proprietary software development and facilitating wider adoption and collection.

Beyond scalability, the VideoCap workflow demonstrates notable efficiency. VideoCap achieved an Annotation Burden Factor of 4.47:1, meaning that each minute of video required approximately 4.5 minutes for viewing, interpretation, and completion of the multi-step survey task.

As shown in Table 2, this burden was lower than the contextual annotation-time estimates reported or derived from prior multimedia annotation workflows, which ranged from 6:1 to 16:1 for surgical procedures, 8:1 to 15:1 for annotation of 100 video concepts, and 14.3:1 for context-aware audiovisual emotion annotation.(15–17)

**Table 2.** Contextual Comparison of Annotation Burden Across Multimedia Annotation Workflows.

| Workflow or Study | Annotation task | Source information | Annotation burden ratio |
| --- | --- | --- | --- |
| VideoCap | Simulated patient-provider video segments with open-ended insights, structured selections, and free-text responses | Mean annotation time: 268.34 seconds; mean video duration: 60 seconds | 4.47:1 |
| Mohamadipanah et al.(15) | Surgical Metrics Project videos with time-stamped task boundaries, maneuvers, instrument use, and tool activity states | Videos were 15-20 minutes; annotation required 2-4 hours per video | 6:1-16:1 |
| Wang et al.(16) | Video annotation across 100 semantic concepts | Annotation of 1 hour of video with 100 concepts typically required 8-15 hours | 8:1-15:1 |
| Clavel et al.(17) | Context-aware audiovisual emotion and situation annotation across pre-segmented sequences | Annotation of 7 hours of pre-segmented audiovisual recordings required approximately 100 hours | 14.3:1 |

**Note:** All annotation burden ratios were calculated by dividing reported annotation time by video duration. For Mohamadipanah et al., the 6:1-16:1 range represents the possible outer range calculated from reported ranges of 15-20-minute videos and 2-4-hour annotation times per video. Ratios are shown for contextual comparison rather than head-to-head performance evaluation because the studies differed in media content, annotation objectives, annotation granularity, and annotator populations.

Furthermore, the process used for tracking completion involves manually downloading survey results periodically and using a Python script to verify the completion of all assigned videos and correct input types. The script then removes completed segments from the CSV file used by the workflow. Further automation could potentially ease the burden of verification.

Future enhancements to our workflow will address key limitations in automation, user experience, and multimodal annotation. Automation could streamline quality assurance by rejecting unsuitable responses based on survey results, ensuring only valid responses proceed to manual review. User experience improvements, such as designing the REDCap survey interface to be more intuitive, with clear and specific instructions, may improve response quality as well as participant experience and engagement. Broader multimodal annotation support will also be important, as many biomedical datasets combine video with audio, transcripts, images, device outputs, or time-stamped metadata. Future deployments could support synchronized presentation of multiple modalities, platform-specific respondent identifiers, automated quality checks, and expert-labeled validation subsets, allowing the same core workflow to support both smaller expert-review studies and larger crowdsourced annotation efforts with minimal modifications.

## Conclusion

In this work, we presented VideoCap, a scalable, secure, and context-aware workflow that leverages Research Electronic Data Capture (REDCap) as a survey platform, Amazon Web Services (AWS) as a backend to track views and generate dynamic survey URLs, and Box, Inc. as a Content Delivery Network (CDN) to facilitate crowdsourced video annotation. Our solution is effective for large datasets while ensuring the safety of distributed content, overcoming many limitations of existing annotation methods.

The design integrates segmented videos with secure hosting, dynamic delivery, and crowdsourced annotation within a unified framework. Additionally, our approach may support broader annotation perspectives by enabling crowdsourced input beyond small expert-only annotator groups.

Our workflow represents an advancement in biomedical informatics by providing a reproducible method for collecting multimedia data annotations that may support downstream machine learning development when paired with appropriate validation. Beyond biomedical applications, this workflow is applicable in various sectors that require precise annotations, including autonomous driving and surveillance.

Looking ahead, the adaptability of our workflow to emerging tools and expanding datasets helps future-proof our solution for any potential multimedia annotation requirements. We encourage adopting and further refining this system to enhance annotation processes and support the growing demands of diverse research and application domains.

## Data Availability

The code and workflow materials used in this study are publicly available at the VideoCap GitHub repository. Aggregate workflow performance metrics supporting the findings of this study are contained within the manuscript. Additional materials are available from the corresponding author upon reasonable request.

https://github.com/kbjohnson-penn/VideoCap

## Code availability

Code utilized across all tools within the workflow, including an importable REDCap survey XML file, is publicly available in the following GitHub repository: https://github.com/kbjohnson-penn/VideoCap.

## CRediT authorship contribution statement

### Basam Alasaly

Conceptualization, methodology, software, formal analysis, data curation, visualization, writing - original draft, writing - review and editing. **Kuk Jin Jang:** Conceptualization, methodology, formal analysis, supervision, writing - original draft, writing - review and editing. **Andrew Zolensky:** Formal analysis, writing - original draft, writing - review and editing. **Sriharsha Mopidevi:** Software, resources, writing - review and editing. **Kevin B. Johnson:** Conceptualization, supervision, project administration, writing - review and editing.

## Acknowledgments

We thank Shawn Ballard, Associate Director for Research Technologies, Clinical Research Collaboration Unit, University of Pennsylvania, for guidance on REDCap survey URL parameterization.

